# Proteomics for the discovery of clinical delirium biomarkers: A systematic review of Major Studies

**DOI:** 10.1101/2022.06.07.22276115

**Authors:** Kwame Wiredu, Edmund Aduse-Poku, Shahzad Shaefi, Scott A Gerber

**Author notes:** **Corresponding author:** Scott A. Gerber, Geisel School of Medicine at Dartmouth, 1 Medical Drive, Lebanon NH.

## Abstract

Delirium represents a significant healthcare burden, diagnosed in over two million elderly Americans each year. In the surgical population, delirium remains the most common complication among elderly patients and is associated with longer hospital stays, higher costs of care, increased mortality and functional impairment. The pathomechanism of disease is poorly understood, with current diagnostic approaches somewhat subjective and arbitrary, and definitive diagnostic biomarkers are currently lacking. Despite the recent interest in delirium research, biomarker discovery for it remains new. Most attempts to discover biomarkers are targeted studies that seek to assess the involvement of one or more members of a focused panel of candidates in delirium. For a more unbiased, systems-biology view, we searched literature from MEDLINE, Cochrane Central, Web of Science, SCOPUS, and Dimensions between 2016 and 2021 for untargeted proteomic discovery studies for biomarkers of delirium conducted on human geriatric subjects. Two reviewers conducted independent review of all search results, and resolved discordance by concensus. From an overall search of 1172 publications, eight peer-reviewed studies met our defined inclusion criteria. The 370 unique peri-operative biomarkers identified in these reports are enriched in pathways involving the activation of the immune system, inflammatory response, and the coagulation cascade. IL-6 was the most commonly identified biomarker. By reviewing the distribution of protein biomarker candidates from these studies, we conclude that a panel of proteins, rather than a single biomarker, would allow for discriminating delirium cases from non-cases. The paucity of hypothesis-generating studies in the peer-reviewed literature also suggests that a systems-biology view of delirium pathomechanisms has yet to fully emerge.

## Introduction

Diagnosed in over 2 million older adults each year, delirium presents a significant healthcare burden in the United States.^1,2^ Delirium is etiologically heterogenous, with many precipitating and predisposing factors.^3-5^ Following surgery, it complicates geriatric hospitalizations with significant functional impairments, longer hospital stays, higher cost of care and increased overall mortality risk.^6-8^ Despite the substantial impact in this demographic, delirium is diagnosed through subjective assessment of a constellation of signs and symptoms within the clinical history, behavioral observation and cognitive assessments.^9^ As a results, commonly used tools such as the confusion assessment method often exhibit inter-rater variability.^10,11^

There is also considerable lack of clarity regarding the pathophysiology of the condition. Given this, it is surprising to note the majority of delirium biomarker research use targeted experiments, where authors study a selected list of biomarkers. The use of targeted strategies for the purposes of discovery, while powerful, is unavoidably biased by the *a priori* knowledge of those biomarkers and the specific focus of the hypothesis under evaluation. Targeted studies may miss as-of-yet unappreciated functional players in a condition as complex as delirium. Furthermore, the biological complexity of commonly used biofluids (such as blood and cerebrospinal fluid) necessitate the use of a measurement platform that is precise and sensitive even for biomarkers of low abundance. Mass spectrometry (MS) remains the gold standard protein discovery platform, although high-throughput platforms such as SOMAscan and proximity extension assays have recentnly been used.^12-14^ Unlike MS, these platforms are semi-targeted and limited in the number of proteins assayable.

The inception of the Network of Investigation of Delirium: Unifying Scientists (NIDUS)^15^ in 2016 to corroborate scientific evidence on delirium has encouraged a more unified nomenclature^16^ and consistency in case identification for the purposes of research. However, only a small proportion of published literature since 2016 has focused on biomarker identification (**Supplemental figure 1**).

The identification of definitive biomarkers of delirium is likely to contribute significantly to our understanding of delirium pathophysiology and to accurately identify cases of this debilitating condition.^17^ Here, we have summarized proteomic contributions in delirium biomarker research in the last six years (2016 – 2021), focusing on untargeted experiments that offer a systems-biology view of the condition. We examine the merits of the different measurement platforms and experimental approaches and have offered perspectives on optimizing sample preparation for the detection of low abundance biomarkers. Lastly, we analyzed the biomarker pool from the published studies for understanding of functional themes that may be at play in the occurrence of delirium.

## Methods

Following the PRISM guidelines,^18^ we searched five databases (MEDLINE, SCOPUS, Central, Web of Science and Dimensions) using the key terms [delirium, acute confusion, acute brain failure] AND [biomarker, biological marker] AND [proteins, proteomics]. Search results were limited to publications written in English and published from 2016 – 2021. EndNote bibliography software version X9.3.3^19^ was used for duplicate removal. All remaining publications were independently reviewed by KW and EAP in a two-stage process. Rayyan freeware, a free web-tool for systematic reviews,^20^ was used to expedite the initial (title and abstract) screening. Secondary screening of remaining publications involved full text review for publications that met the inclusion criteria of (1) untargeted proteomic profiling, (2) for biomarkers of delirium, (3) conducted on human geriatric subjects.

Discordance was resolved by both KW and EAP through consensus. Biomarker data from included studies were collected from the online archives of the respective study journals. Here, a biomarker was deemed differentially abundant if the degree of fold-change relative to study controls of at least ±1 was specified by authors at a significance level of ≤ 0.05. Analysis for over-represented functional themes (ie biological processes, cellular components and molecular functions) for performed using the gene ontology (GO) database from UniprotKB (last accessed 05/18/2022). Data retrieval and visualizations were achieved using R environment for statistical computing, v4.1.1.^21^

## Results

We identified a total of 1,172 publications from 5 database searches (MEDLINE, SCOPUS, Central, Web of Science and Dimensions). The majority of articles excluded during initial screening were review articles, non-biomarker articles, poster abstracts, meeting proceedings, editorial and comments. Full text review was performed on 280 articles, many of which were either targeted biomarker studies on delirium, animal studies or involved non-protein delirium biomarkers such as brain imaging. Unique in these exclusions was one study on delirium in children. **Figure 1** is a flowchart of the screening steps and exclusion criteria, leading to the final eight peer-reviewed original studies summarized in **Table 1**. Of the eight studies, five were conducted in North America ^22-26^ and one each in Asia ^27^, Europe ^28^ and Sweden ^29^.

**Figure 1:**
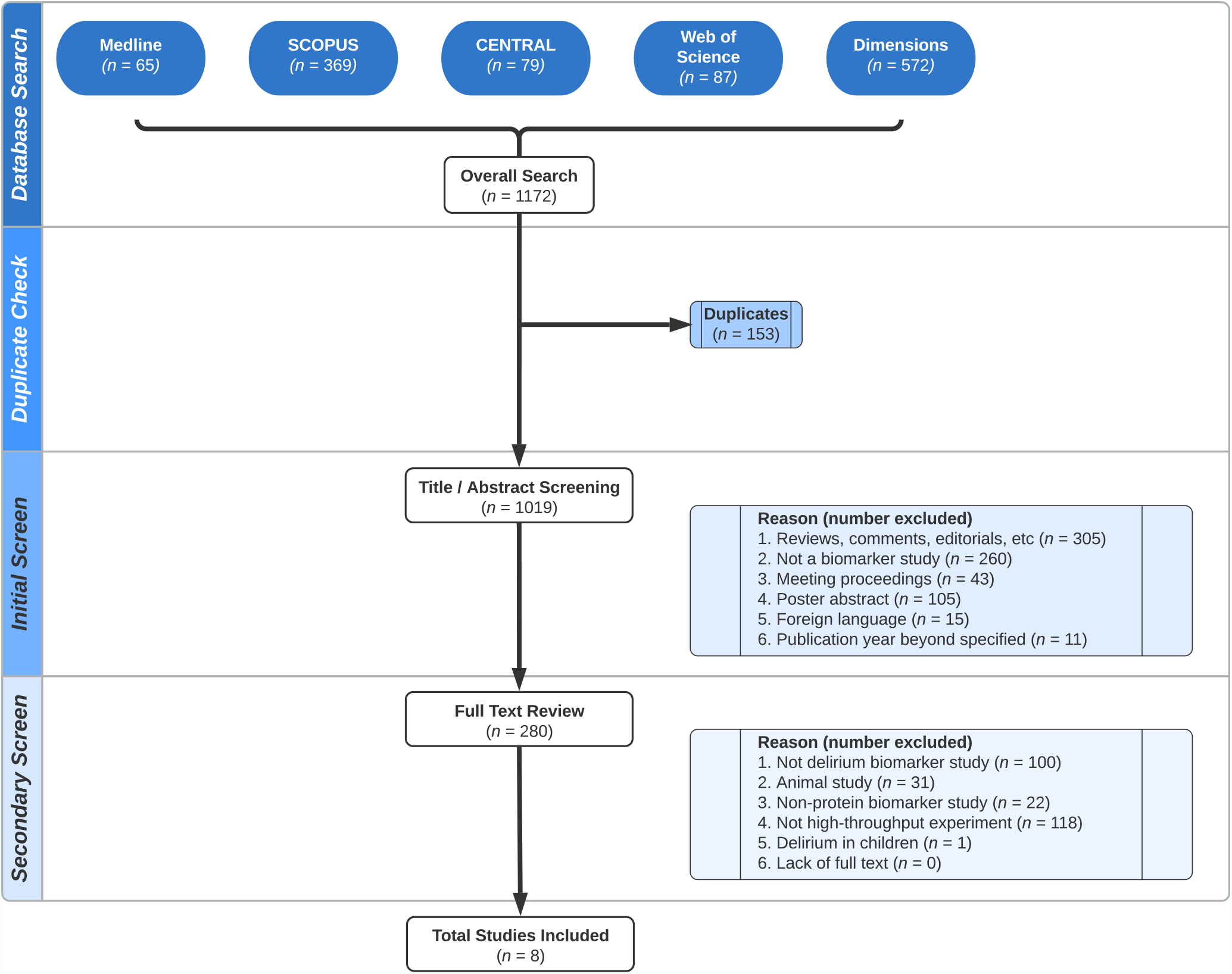
Literature search and screening: PRISMA flow chart highlights the step-by-step process involved in the selection of the final 8 studies summarized in this review.

### Study Design, Patient Selection and Choice of Controls

All but two studies had a nested, case-control design.^28,29^ Overall, the age of delirium cases averaged 73.3 years (**Table 1**). Samples from a total number of 484 subjects (cases and controls) were used for biomarker discovery alone, although there is likely an overlap in subjects selected from the same parent study (i.e., the SAGES study or the MINDDS trial). Except for one study which profiled biofluids from non-surgical patients^28^, eligible subjects were all surgical patients, who underwent either cardiac^25,26^ or non-cardiac procedures.^22-24,27,29^ Patients’ comorbidity score, either with the Charlson index or PROMIS, was established in all but for two studies^28,29^, although van Ton, Verbeek, Alkema, Pickkers, Abdo ^28^ indicated that hypertension, diabetes, immunosuppression and cerebrovascular disorders were common in the selected cohort.

Delirium cases were identified with either the Confusion Assessment Method (CAM), the Chart-based Delirium Identification Instrument (CHART-DEL) or the Diagnostic and Statistical Manual of Mental Disorders (DSM-V) (**Table 1**). Subjects from the MINDDS trial^25,26^ received twice daily post-operative assessments for delirium occurrence, compared with once daily assessments for the SAGES cohort^22-24^.

**Table 2.1:**
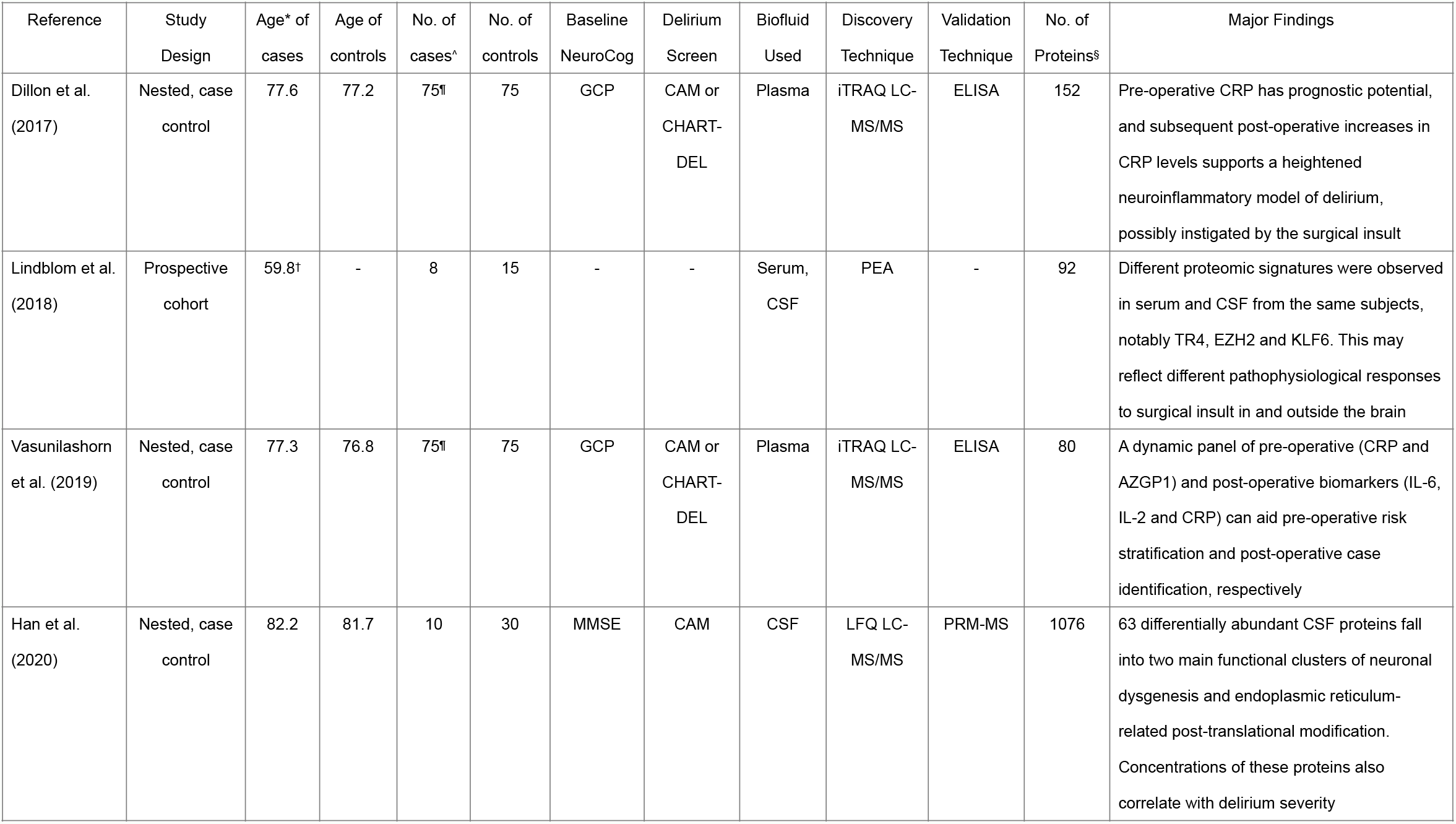
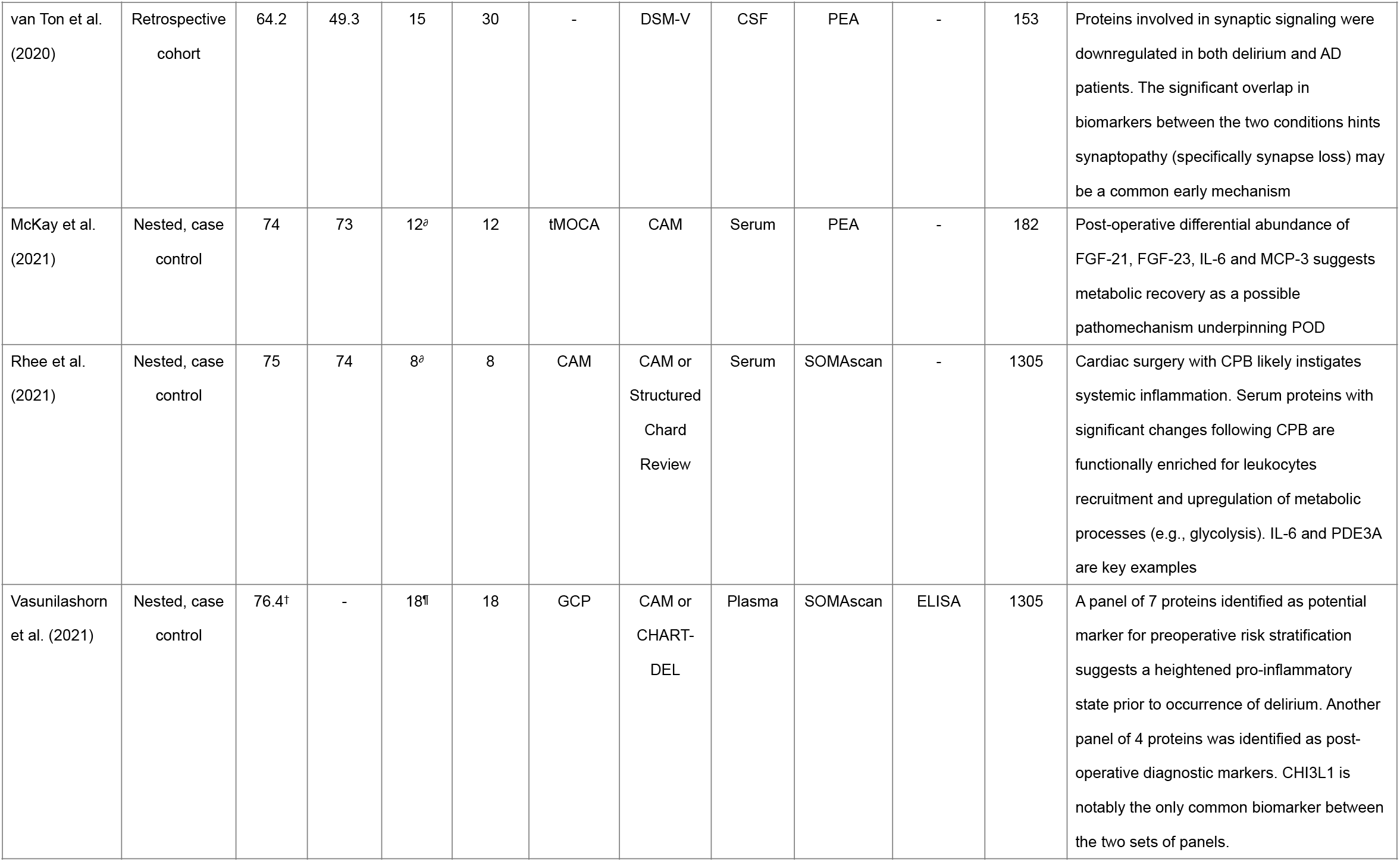
Summary of included studies

Baseline neurocognition of study participants was established in seven studies (**Table 1**). Neither the case identification method nor baseline neurocognition approach was specified by Lindblom, Shen, Axen, Landegren, Kamali-Moghaddam, Thelin ^29^. All eight studies used non-delirium controls to establish a statistical baseline, with some variations in the choice of controls. Controls were age- and sex-matched in five studies^22-26^, although the SAGES cohort included baseline cognitive performance as an additional matching parameter. Han, Chen, Song, Yuan, Li, Zhou, Liu, Han, Mi, Li, Wang, Zhong, Zhou, Guo ^27^ selected controls that matched to cases by age and by mini-mental state examination (MMSE). In the study by van Ton, Verbeek, Alkema, Pickkers, Abdo ^28^, two groups of controls to the post-infectious delirium cases were selected: healthy controls and controls with neurocognitive impairments other than delirium. Similarly, Lindblom, Shen, Axen, Landegren, Kamali-Moghaddam, Thelin ^29^ selected surgical patients who suffered other neurological injuries but without the diagnosis of delirium.

### Sample Preparation and Proteomic Techniques

The source of proteins for biomarker discovery included peripheral blood (five studies), cerebrospinal fluid (CSF; two studies) and both blood and CSF (one study) (**Table 1**). When CSF was used, lumbar puncture samples were collected only once. This contrasts with the serial collection of samples for the blood-based studies. Three of the six blood-based studies used plasma and the remaining three used serum. Of note, only the plasma-based studies ^22-24^ documented sample immunodepletion, specifically by using affinity-based depletion columns to remove the 14 most abundant proteins in an effort to detect proteins of lower abundance. Three studies used mass spectrometry (MS) as the analytical approach, three studies used proximity extension assays (PEA), and the remaining two used SOMAscan technology. Two studies attempted sample multiplexing with isobaric labelling^22,23^. There was, however, no mention of pre-analytical sample fractionation to further reduce sample complexity in any of the studies.

### Proteomic Biomarkers

In the union of all eight studies, 446 unique proteins were identified as candidate biomarkers for delirium. Of these biomarkers, 370 were identified peri-operatively. **Figure 2** and **Supplemental Figure 2** illustrate the contribution of each study to the total pool of candidate biomarkers, and where biomarkers overlap between studies. Overall, Vasunilashorn, Dillon, Chan, Fong, Joseph, Tripp, Xie, Ngo, Lee, Elias ^24^ reported the largest number of differentially abundant proteins (*n* = 128) between cases and controls. Interleukin-6 (IL-6) was the most commonly identified, differentially abundant protein among the studies ^24-26,28,29^. Complement component C9, antithrombin-III (SERPINC1), the cytokine fractalkine (CX3CL1) and chitinase-3-like protein 1 (CHI3L1) are notable biomarkers that were found in three or more studies. Except for the studies done in the SAGES cohort^22-24^, very few of the remaining proteins overlap between studies.

**Figure 2:**
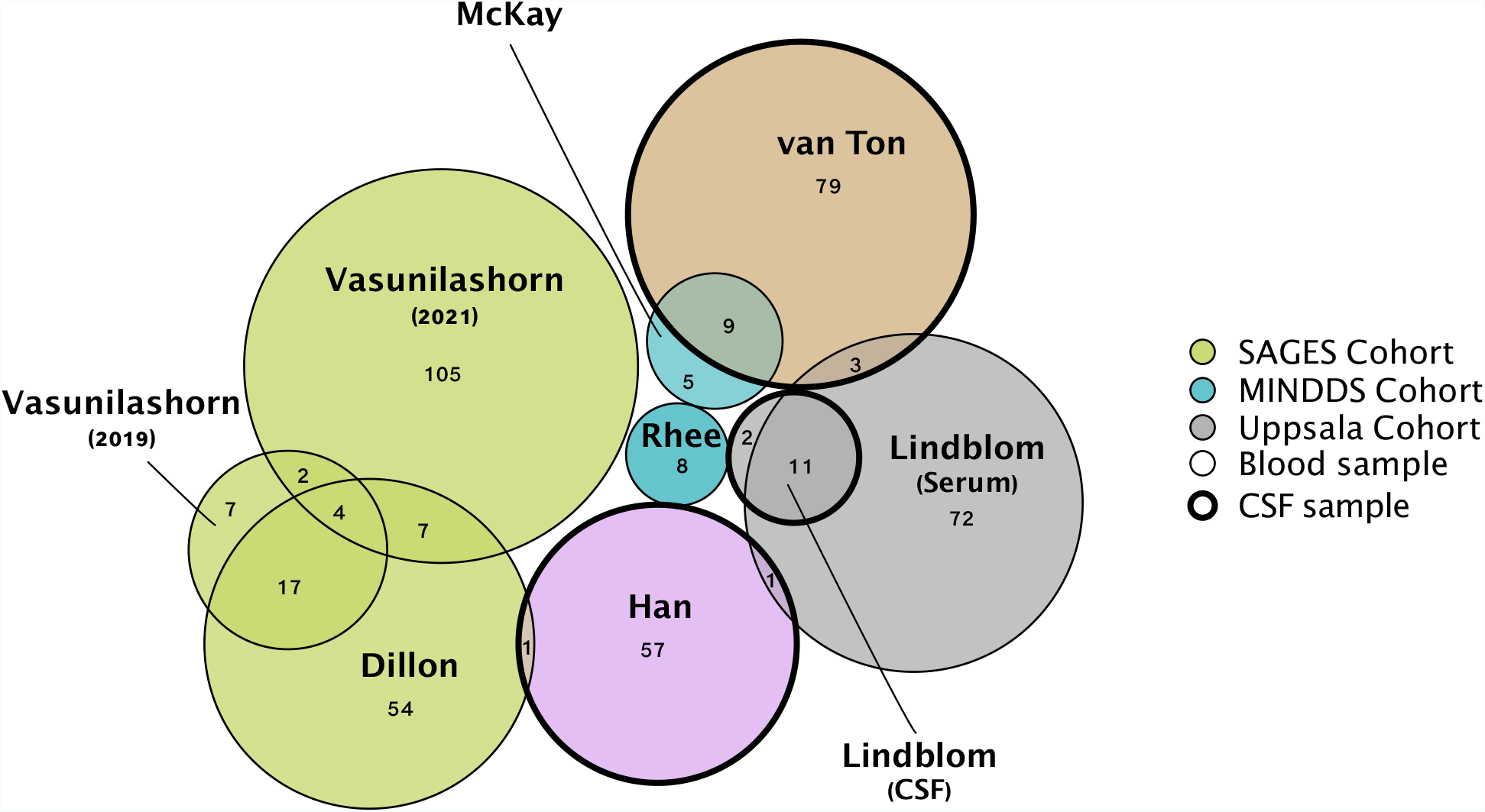
Modified Venn (Euler) diagram showing the intersecting biomarkers between the 8 studies. Size of circle is proportional to number of biomarkers. Colors represent studies from the same cohort, with likely overlap in the subjects selected for proteomic profiling. Thickness of circle outline indicates the type of biofluid used. NB: Intersections between 4 or more studies are not visualized here.

Functional analysis on the peri-operative biomarker pool of 370 proteins for enriched biological processes suggests a systemic response of widespread activation and dysregulation of proteins involved in immunological reactions, inflammatory responses, and the coagulation cascade (**Figures 3A, 3D**). Furthermore, subcellular ontology annotation reveals the extracellular region as the predominant native location of these dysregulated proteins, enriched in signaling and cytokine activity (**Figures 3C, 3D**).

**Figure 3:**
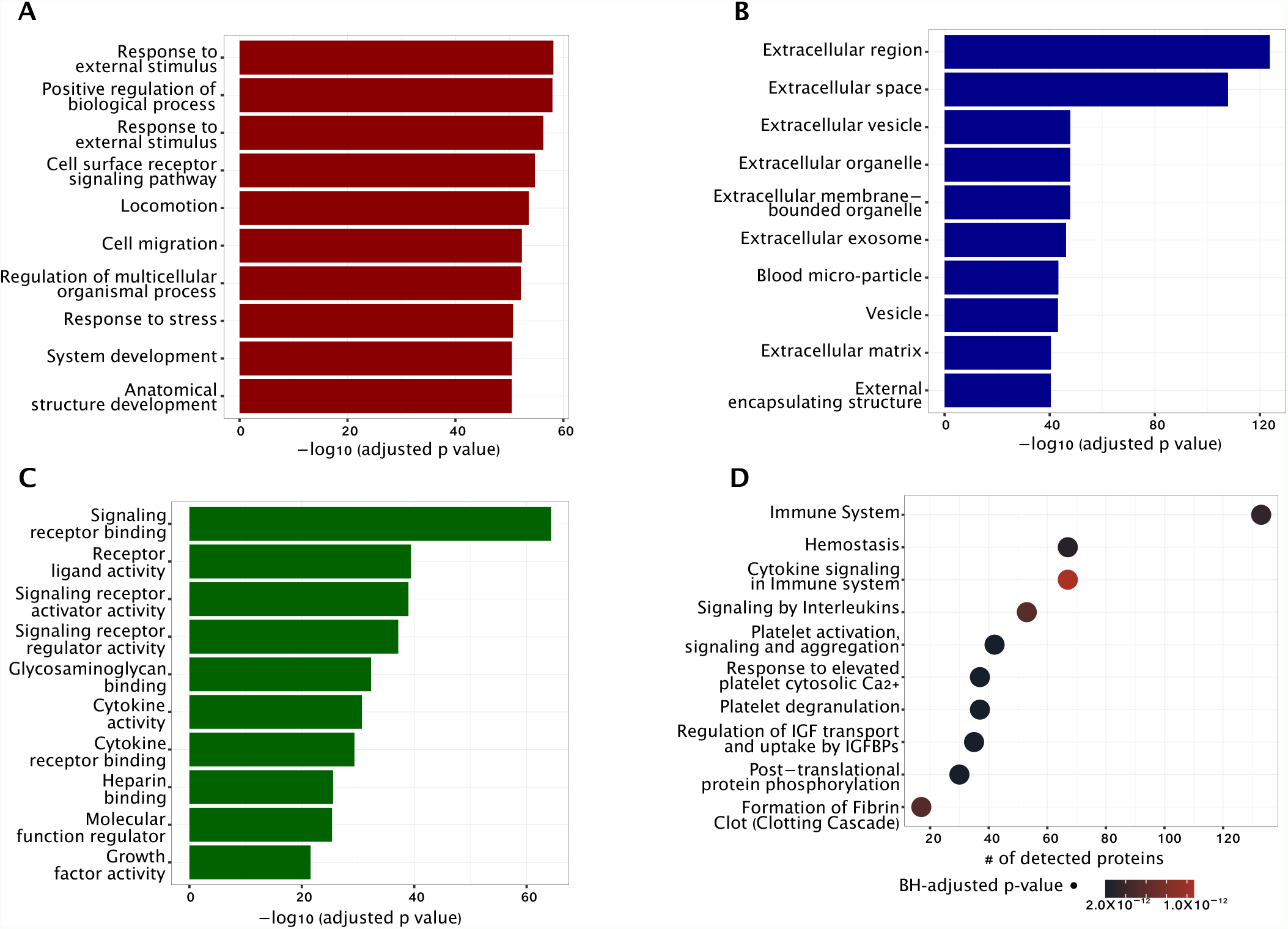
Functional analysis of the biomarker pool showing the top 10 GO terms with regards to (A) biological processes, (B) cellular component, and (C) molecular functions. The number of proteins involved in each of the major functional classes in the biomarker pool are shown in (D).

### Validation Approaches

All three mass spectrometry-based studies and one SOMAscan-based study performed protein verification and validation on candidate biomarkers identified by high throughput methods (**Table 1**). Of these studies, three used ELISA, an affinity-based approach^22-24^ and the remaining study used a MS-based approach, specifically parallel reaction monitoring (PRM).^27^ The choice of biomarkers that were validated is however varied. Of the 63 differentially abundant proteins in the study by Han, Chen, Song, Yuan, Li, Zhou, Liu, Han, Mi, Li, Wang, Zhong, Zhou, Guo ^27^, 20 were selected for validation by PRM based on a minimum number of peptides and transitions set by the authors. Of the remaining studies, the choice to validate CRP, SERPINA3, AZGP1 and CHI3L1 was based, partly, on the consistency of their identification in various samples, the availability of a commercial antibody and a series of binomial, signed rank and Student *t-*tests.

## Discussion

We have presented an in-depth review of clinical proteomic contributions over the preceding six complete years that offered an unbiased systems-biology view of delirium. As consistency in case identification and unified nomenclature is necessary to make comparisons between studies, we began our literature search from 2016. We observed that this is also the year NIDUS was established,^15^ and interest in delirium research has seen a steady increase since that point. We focused on studies that measured delirium biomarkers using a discovery-based approach in the human geriatric population. The total of eight studies that met our criteria signify the paucity of literature that offers a systems-biology view of delirium in this demographic. Nonetheless, the large proportion of search results that were excluded as grants, conference abstracts, meeting proceedings, and posters suggest a growing interest in research on delirium diagnostics.

We aggregated a total of 446 biomarkers that are differentially dysregulated in human patients with delirium across eight studies. It is worth noting that subjects in one study^28^ developed delirium after an infectious process. Functional analysis of the 370 peri-operative pool of biomarkers suggests a widespread activation of immunological reactions, inflammatory responses, and the coagulation cascade. We focused functional analyses only on biomarkers discovered peri-operatively, given that infectious delirium may involve a different pathophysiological process^28,30-32^, although analyses of all 446 biomarkers did not reveal any functional differences. Given that IL-2, C-X-C motif chemokine 11 and C-C motif chemokine 13 ^23-25^, among others, were elevated preoperatively, it is equally likely that a heightened pre-operative inflammatory state increased the risk of delirium, although functional studies would be required to rigorously test this hypothesis. This observation is consistent with prevailing knowledge that phenotypic delirium is a culmination of multiple predisposing and precipitating factors^5,33,34^, and predictive biomarkers may be more beneficial in certain risk groups than others.

At the cellular level, the biomarker pool is enriched in signaling and cytokine activity, predominantly in the extracellular domain. While there are many extracellular domain-containing proteins that do not localize to synapses, this observation may signify the possibility of altered synaptic functioning in the context of delirium. Synaptic dysfunction is an early event in Alzheimer’s disease^35^, and many researchers have suggested similar findings as a common pathophysiological starting point in the continuum of neurocognitive disorders, of which delirium and AD are a part^36-38^.

Interleukin-6 remains one of the most consistently identified proteins among delirium patients. In well-functioning older patients, IL-6 is found to be prospectively associated with cognitive decline^39-41^. IL-6 is also part of the core panel of frailty biomarkers ^42^, and has recently been suggested by Gómez-Rubio, Trapero, Cauli, Buigues ^43^ as a useful biomarker for monitoring treatment in frail individuals. The inflammatory role of IL-6 and its associations with aforementioned predisposing triggers further emphasize the neuro-inflammatory model of delirium.

Notable in the 24 overlapping proteins between Dillon, Vasunilashorn, Ngo, Otu, Inouye, Jones, Alsop, Kuchel, Metzger, Arnold, Marcantonio, Libermann ^22^ and Vasunilashorn, Ngo, Chan, Zhou, Dillon, Otu, Inouye, Wyrobnik, Kuchel, McElhaney, Xie, Alsop, Jones, Libermann, Marcantonio ^23^ is CRP, an acute phase reactant and a non-specific marker of inflammation, infection and tissue injury^44^. Many authors have suggested elevated CRP levels to be associated with a higher risk of delirium occurrence^45-50^, and could likely be used to monitor the clinical course of delirium ^51^. In a recent meta-analysis of 54 observational studies, Liu, Yu, Zhu ^52^ hinted that CRP may be a more specific marker of post-operative delirium (POD) than post-operative cognitive dysfunction (POCD). The clinical relevance of this specificity remains unclear, given that a lower baseline cognitive reserve is a precipitating factor for both POD and POCD^53^.

It is well documented that elevated total cholesterol and LDL correspond to increased neuritic plaque density in Alzheimer’s disease ^54,55^. One overlapping protein, apolipoprotein A-IV (APOA4) is a component of chylomicrons and HDL, synthesized mainly in the intestine and secreted into plasma^56^. Although there is some tenuous evidence of an association with cognitive impairment and Alzheimer’s disease^57,58^, data on APOA4 is scarce and there has yet to be a formal interrogation of its association in delirium. This holds true for many proteins in this union of 446 biomarkers.

Furthermore, it is also unclear how the modest degree of overlap in candidate biomarkers between the eight studies reflect differences in the biofluids used or the analytical strategies used to identify the biomarkers. While MS relies on peptide spectrum matches for protein identification, PEA (Olink Proteomics, Uppsala, Sweden) is an oligonucleotide-based immunoassay that combines quantitative real-time PCR with high-throughput quantification. SOMAscan (SomaLogic, Inc, Boulder, CO), on the other hand, uses aptamers to bind specific molecular targets. These affinity technologies have recently gained attention in plasma proteomics as they are cost-effective, require less expertise, much smaller sample volumes and can quantify a little over 1000 human plasma proteins. In fact, the number of recent original publications on plasma proteomics that use PEA outnumber those that present MS-based approaches^13^. Despite reports of comparable reproducibility and complementarity of PEA and SOMAscan to MS, a recent study comparing PEA to MS-based protein profiling revealed a similar modest degree of overlapping proteins as found in this review^13^. Of the 14 available PEA panels, van Ton, Verbeek, Alkema, Pickkers, Abdo ^28^ and McKay, Rhee, Colon, Adelsberger, Turco, Mueller, Qu, Akeju ^25^ used the two that predominantly assay neural and inflammatory markers. It is therefore not surprising that there are nine overlapping biomarkers between these two studies. The SOMAscan-based studies had only one biomarker in common, namely IL-6. Given that these affinity-based platforms are semi-targeted and predominantly assay the low abundance plasma proteome, it is our thinking that the strengths of all three analytical techniques could be viewed as complementary, offering a deeper view into the plasma proteome together than each would separately.

Three of the six blood-based studies used serum which is qualitatively and quantitatively different from plasma. Removal of clotting factors (largely fibrinogen) results in a 3 – 4% lower protein concentration in serum relative to plasma^59,60^, and may also lead to removal of proteins with specific (or non-specific) interactions with fibrin in a manner that is unpredictable. The Human Proteome Organization (HUPO) therefore endorses the use of plasma, citing a lower degree of *ex vivo* degradation and recommends that citrate or EDTA be used for anticoagulation over heparin^61^. As the sample choice should be tailored to the specific biomarker needs and the biomarker landscape of delirium is still in its infancy stages, it would be preferable that the biofluid used, their collection and sample preparation protocols permit the study of the entire plasma proteome.

Serial collection of samples in the blood-based studies allowed for the determination of temporal associations of proteomic changes with the occurrence of delirium. In all but one of the blood-based studies^29^, a minimum of two samples were collected for each study participant: at baseline (pre-operative) and on post-operative day one. This is in sharp contrast to the CSF-based studies which were limited by the one-time sample collection by lumbar puncture. CSF is the proximal biofluid of choice with a greater likelihood of reflecting the immediate proteomic changes in the brain. Unfortunately, CSF access is severely limited by the invasive nature of the sampling technique. Furthermore, it is hypothesized that the relatively higher permeability of the blood brain barrier of the elderly brain makes it possible for proteomic changes in CSF to be detected in plasma.

Immunodepletion is generally thought to be beneficial because of the wide dynamic range (≂10^10^) of protein abundances in plasma and CSF, dominated by a handful of highly abundant proteins such as albumin and immunoglobulins. This makes identifying and quantifying proteins of lower abundance otherwise difficult. Three of the eight studies reported use of immunodepletion on the samples prior to analysis. Given that PEA and SOMAscan are semi-targeted, it is unclear if immunodepletion is a necessary pre-analytical step. The extent to which the use immunodepletion, or otherwise, affected the identification of candidate delirium biomarkers is unclear. In addition, fractionation strategies such as ion exchange chromatography significantly reduce sample complexity and increase the depth of proteome coverage, especially when searching for low abundance plasma proteins. With the collective results from all eight studies indicating possible neuro-inflammatory process(es) to play a prominent role in delirium pathogenesis, candidates-biomarkers of delirium are likely to be in the low abundance proteome, which makes use of immunodepletion and sample fractionation an important consideration in the experimental design. Clearly, the relative advantages and disadvantages of each sample type and the approach to sample preparation will continue to play a significant role in the design of future studies to identify protein biomarkers in delirium and neurocognitive disorders at large.

Integrating results from eight different studies was not without challenges, one of which was the use of different delirium diagnostic algorithms. Even within the same study, Dillon, Vasunilashorn, Ngo, Otu, Inouye, Jones, Alsop, Kuchel, Metzger, Arnold, Marcantonio, Libermann ^22^ employed both CAM and chart review for delirium case identifications. Further, the inclusion of subsyndromal delirium (SSD) cases together with delirium cases in the studies by Dillon, Vasunilashorn, Ngo, Otu, Inouye, Jones, Alsop, Kuchel, Metzger, Arnold, Marcantonio, Libermann ^22^, Vasunilashorn, Ngo, Chan, Zhou, Dillon, Otu, Inouye, Wyrobnik, Kuchel, McElhaney, Xie, Alsop, Jones, Libermann, Marcantonio ^23^ and Vasunilashorn, Dillon, Chan, Fong, Joseph, Tripp, Xie, Ngo, Lee, Elias ^24^ may further complicate data interpretation in the context of other studies. In addition, none of the studies formally screened for depression among the study subjects. Lastly, because different studies reported protein names using either accession number, gene name or Uniprot protein annotations, we had to manually convert all names to a common nomenclature to allow for comparisons and functional analyses.

Lastly, nested case control is an adequate study design for biomarker studies. It is however limited in precision, inferential conclusions and power due to sampling of controls. Only associations, and not causal inferences, can be concluded from nested designs, even after adjusting for most confounding variables^62,63^. The choice of controls to establish a statistical baseline plays a significant role in subsequent differential abundance analyses. In the study by van Ton, Verbeek, Alkema, Pickkers, Abdo ^28^, controls were significantly younger than delirium cases (49.3 versus 64.2 years). Nonetheless, this is the only study in which authors excluded control subjects with a known acute or chronic systemic inflammation.

## Conclusion

The urgent need for diagnostic and predictive biomarkers of delirium is important not only to correctly identify cases, but also for pre-operative risk stratification and for follow-up on possible long-term neurocognitive sequelae. Interest in delirium research has seen a steady rise since the inception of NIDUS in 2016. Nonetheless, delirium biomarker research appears to be just emerging. There are only a handful of studies that offer a systems-biology view of delirium from human patient samples. For diagnostic purposes, it appears likely that a panel of biomarkers, rather than a single biomarker, has potential for discriminating delirium cases from non-cases. Collectively, biomarkers from these studies suggest an immunological and inflammatory response following surgical insult, enriched in cytokine and signaling activity in the extracellular space. Further studies are warranted to support this observation. With a greater focus on the low-abundance plasma proteome, complementary use of MS and PEA may yield a deeper plasma proteome profiling.

## Data Availability

All data produced in the present study are available upon reasonable request to the authors

## Acknowledgment

We acknowledge Elaina Vitale, a Research and Education Librarian at the Dartmouth College library for their input on the search strategy. Some of the funding for this work was supplied by the Burroughs Wellcome training grant to KW, the National Institutes of Health (R01 GM122846) to SAG, and K08 GM134220 and R03 AG060179 to SS.

## Author contributions

KW conceptualized the study, collected and analyzed data, and drafted the manuscript. EAP assisted with data collection and review of search results. Both SS and SAG guided study conceptualization, data collection and analysis, revised the manuscript and provided funding.

This review is part of an ongoing PhD thesis project at the Geisel School of Medicine at Dartmouth.

## Data accessibility statement

Biomarker data is available as supplemental material. Analytic codes for data wrangling, functional analyses and visualization can be made available upon request.

## Captions

**Supplemental figure 1:**
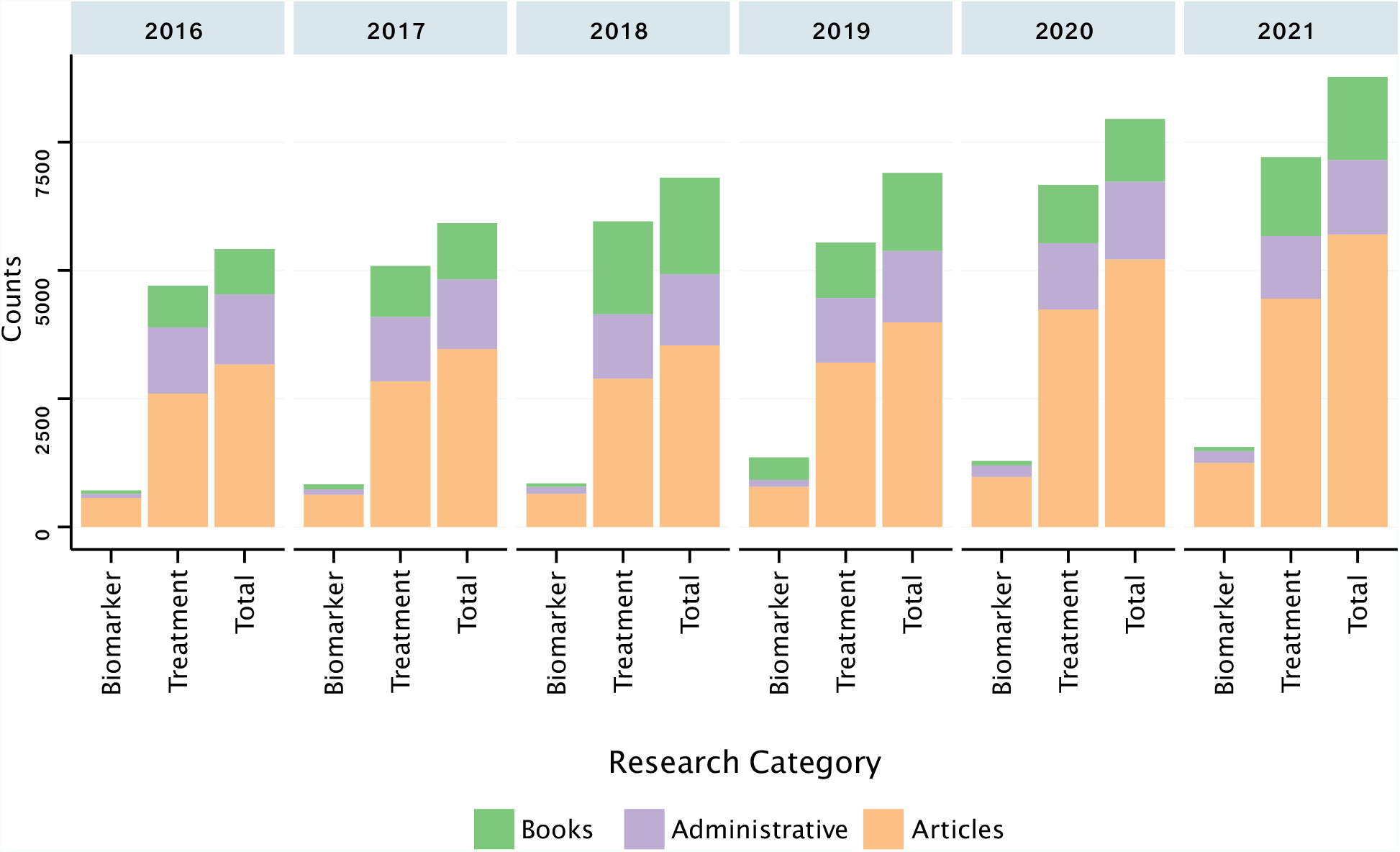
Counts of all published documents on delirium between 2016 and 2021, highlighting documents involved with delirium biomarkers only, or delirium treatment and prevention. Documents described as articles include peer-reviewed original research, study protocols, preprints, poster abstracts, monographs, conference proceedings and editorials and opinions. Administrative documents include grants, patents, clinical trials and policy documents. (Source of data: Dimensions.ai^64^, downloaded on 03/25/2022)

**Supplemental figure 2:**
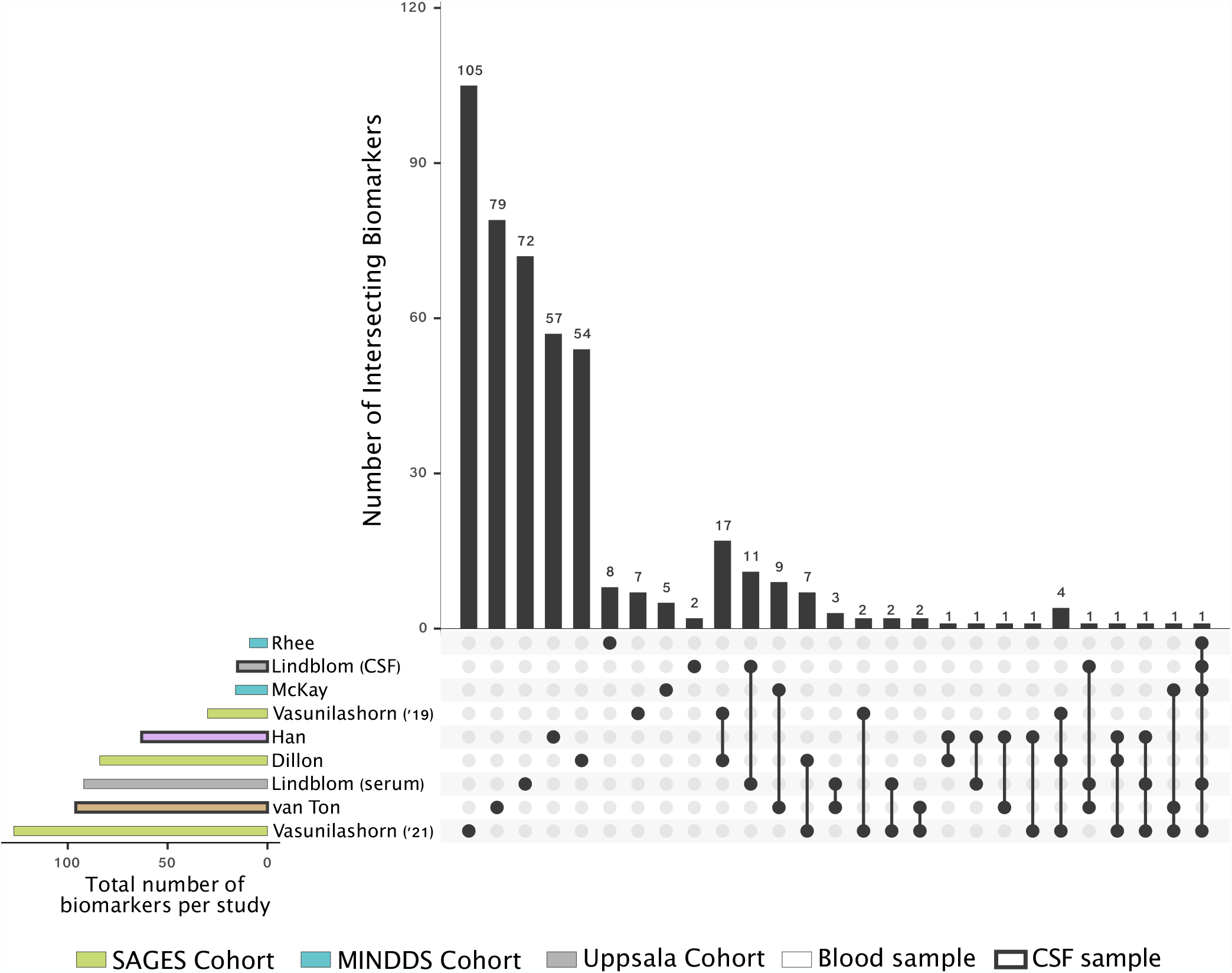
UpSet chart showing the contributions of each study to the total pool of 446 unique biomarkers, and all the intersecting sets of proteins that could not be illustrated in figure 2 (main). Same colors represent studies from the same cohort, with likely overlap in the subjects selected for proteomic profiling. Thickness of rectangle outline indicates the type of biofluid used.

PRISMA 2020 Checklist
| Section and Topic | Item # | Checklist item | Location where item is reported |
| --- | --- | --- | --- |
| **TITLE** |  |  |  |
| Title | 1 | Identify the report as a systematic review. | pp 1 |
| **ABSTRACT** |  |  |  |
| Abstract | 2 | See the PRISMA 2020 for Abstracts checklist. | pp 2 |
| **INTRODUCTION** |  |  |  |
| Rationale | 3 | Describe the rationale for the review in the context of existing knowledge. | pp 3 – 4 |
| Objectives | 4 | Provide an explicit statement of the objective(s) or question(s) the review addresses. | pp 3 – 4 |
| **METHODS** |  |  |  |
| Eligibility criteria | 5 | Specify the inclusion and exclusion criteria for the review and how studies were grouped for the syntheses. | pp 4 |
| Information sources | 6 | Specify all databases, registers, websites, organisations, reference lists and other sources searched or consulted to identify studies. Specify the date when each source was last searched or consulted. | pp 4 |
| Search strategy | 7 | Present the full search strategies for all databases, registers and websites, including any filters and limits used. | Figure 1, supplemental table 1 |
| Selection process | 8 | Specify the methods used to decide whether a study met the inclusion criteria of the review, including how many reviewers screened each record and each report retrieved, whether they worked independently, and if applicable, details of automation tools used in the process. | pp4, figure 1, supplemental table 1 |
| Data collection process | 9 | Specify the methods used to collect data from reports, including how many reviewers collected data from each report, whether they worked independently, any processes for obtaining or confirming data from study investigators, and if applicable, details of automation tools used in the process. | pp 4-5 |
| Data items | 10a | List and define all outcomes for which data were sought. Specify whether all results that were compatible with each outcome domain in each study were sought (e.g. for all measures, time points, analyses), and if not, the methods used to decide which results to collect. | pp 4, figure 1 |
|  | 10b | List and define all other variables for which data were sought (e.g. participant and intervention characteristics, funding sources). Describe any assumptions made about any missing or unclear information. | Table 1 |
| Study risk of bias assessment | 11 | Specify the methods used to assess risk of bias in the included studies, including details of the tool(s) used, how many reviewers assessed each study and whether they worked independently, and if applicable, details of automation tools used in the process. | pp 4-5 |
| Effect measures | 12 | Specify for each outcome the effect measure(s) (e.g. risk ratio, mean difference) used in the synthesis or presentation of results. | pp 4-5 |
| Synthesis methods | 13a | Describe the processes used to decide which studies were eligible for each synthesis (e.g. tabulating the study intervention characteristics and comparing against the planned groups for each synthesis (item #5)). | pp4, table 1 |
|  | 13b | Describe any methods required to prepare the data for presentation or synthesis, such as handling of missing summary statistics, or data conversions. | pp 4-5 |
|  | 13c | Describe any methods used to tabulate or visually display results of individual studies and syntheses. | pp 4-5 |

PRISMA 2020 Checklist
| Section and Topic | Item # | Checklist item | Location where item is reported |
| --- | --- | --- | --- |
|  | 13d | Describe any methods used to synthesize results and provide a rationale for the choice(s). If meta-analysis was performed, describe the model(s), method(s) to identify the presence and extent of statistical heterogeneity, and software package(s) used. | pp 4-5 |
|  | 13e | Describe any methods used to explore possible causes of heterogeneity among study results (e.g. subgroup analysis, meta-regression). | - |
|  | 13f | Describe any sensitivity analyses conducted to assess robustness of the synthesized results. | - |
| Reporting bias assessment | 14 | Describe any methods used to assess risk of bias due to missing results in a synthesis (arising from reporting biases). | pp 4-5 |
| Certainty assessment | 15 | Describe any methods used to assess certainty (or confidence) in the body of evidence for an outcome. | pp 4-5 |
| **RESULTS** |  |  |  |
| Study selection | 16a | Describe the results of the search and selection process, from the number of records identified in the search to the number of studies included in the review, ideally using a flow diagram. | pp 5-9, figure 1 |
|  | 16b | Cite studies that might appear to meet the inclusion criteria, but which were excluded, and explain why they were excluded. | pp 5-9 |
| Study characteristics | 17 | Cite each included study and present its characteristics. | pp 5-9, table 1 |
| Risk of bias in studies | 18 | Present assessments of risk of bias for each included study. | - |
| Results of individual studies | 19 | For all outcomes, present, for each study: (a) summary statistics for each group (where appropriate) and (b) an effect estimate and its precision (e.g. confidence/credible interval), ideally using structured tables or plots. | pp 5-9, table 1 |
| Results of syntheses | 20a | For each synthesis, briefly summarise the characteristics and risk of bias among contributing studies. | table 1 |
|  | 20b | Present results of all statistical syntheses conducted. If meta-analysis was done, present for each the summary estimate and its precision (e.g. confidence/credible interval) and measures of statistical heterogeneity. If comparing groups, describe the direction of the effect. | Figures 2, 3, supplemental fig 2 |
|  | 20c | Present results of all investigations of possible causes of heterogeneity among study results. | - |
|  | 20d | Present results of all sensitivity analyses conducted to assess the robustness of the synthesized results. | pp 5-9 |
| Reporting biases | 21 | Present assessments of risk of bias due to missing results (arising from reporting biases) for each synthesis assessed. | - |
| Certainty of evidence | 22 | Present assessments of certainty (or confidence) in the body of evidence for each outcome assessed. | pp 5-9 |
| **DISCUSSION** |  |  |  |
| Discussion | 23a | Provide a general interpretation of the results in the context of other evidence. | pp 9-16 |
|  | 23b | Discuss any limitations of the evidence included in the review. | pp 9-16 |
|  | 23c | Discuss any limitations of the review processes used. | pp 9-16 |

PRISMA 2020 Checklist
| Section and Topic | Item # | Checklist item | Location where item is reported |
| --- | --- | --- | --- |
|  | 23d | Discuss implications of the results for practice, policy, and future research. | pp 9-16 |
| **OTHER INFORMATION** |  |  |  |
| Registration and protocol | 24a | Provide registration information for the review, including register name and registration number, or state that the review was not registered. | Not registered |
|  | 24b | Indicate where the review protocol can be accessed, or state that a protocol was not prepared. | Not prepared |
|  | 24c | Describe and explain any amendments to information provided at registration or in the protocol. | - |
| Support | 25 | Describe sources of financial or non-financial support for the review, and the role of the funders or sponsors in the review. | pp 17 |
| Competing interests | 26 | Declare any competing interests of review authors. | pp 17 |
| Availability of data, code and other materials | 27 | Report which of the following are publicly available and where they can be found: template data collection forms; data extracted from included studies; data used for all analyses; analytic code; any other materials used in the review. | Biomarker data. Analytic codes available upon request |

From: Page MJ, McKenzie JE, Bossuyt PM, Boutron I, Hoffmann TC, Mulrow CD, et al. The PRISMA 2020 statement: an updated guideline for reporting systematic reviews. BMJ 2021;372:n71. doi: 10.1136/bmj.n71
For more information, visit: http://www.prisma-statement.org/

